# Biological aging clock from routine clinical and anthropometric measurements in diverse populations

**DOI:** 10.64898/2026.05.20.26353724

**Authors:** Alejandro Mejia-Garcia, Chen-Yang Su, Thomas M. Zheng, Hsuan Megan Tsao, Arthur Richard, Dylan Hamitouche, Satoshi Yoshiji, Vincent Mooser, Guillaume Lettre, Adil Harroud, Sirui Zhou

## Abstract

Aging is accompanied by a progressive decline in physiological function that contributes to chronic disease development. Biological clocks estimated from high-dimensional clinical and biological measurements may provide more granular tracking of the aging processes. Current biological clocks, however, have limited cross-ancestry generalizability and clinical applicability. Here, we developed a multi-ancestry biological clock (ClinBAG) using 22 routine blood and anthropometric biomarkers in 14,328 age-and sex-balanced individuals from the All of Us Research Program. We tested the association of ClinBAG with 434 traits and evaluated its ability to predict incident disease in 152,733 non-overlapping individuals. We also conducted genome-wide association studies in European (N=74,675), African (N=22,315), and Admixed American ancestry individuals (N=19,940). Among 190 neurological phenotypes, elevated ClinBAG was associated with cognitive decline, increased incidence of dementia (HR=1.020, p=1.6x10^-5^) and Parkinson’s disease (HR=1.014, p=0.023), and decreased risk of migraine (HR=0.991, p=8.7x10^-4^). We also identified common (*NPRL3*) and ancestry-specific genetic loci (*HBB* in African-ancestry and *FADS1/FADS2* in European-ancestry) for ClinBAG. Single-cell enrichment revealed that ClinBAG-associated genes are overexpressed in double-negative (DN) T cells in an age-dependent manner. This study presents a clinically applicable multi-ancestry biological age clock predicting neurological disease risk. Our findings also uncover population-specific genetic drivers, particularly involving erythropoiesis and DN T-cell-mediated neuroinflammation.

## Introduction

Aging is accompanied by a progressive decline in physiological function that contributes to the development of chronic diseases. Most common neurological conditions, such as stroke, dementia, and Parkinson’s disease, show a strong age-related increase in incidence (1). Even for diseases that may not appear to be aging-related, such as multiple sclerosis (MS), which often begins in young adulthood, age remains the strongest determinant of outcome (2). Thus, aging is central not only to disease onset but also to disease progression and prognosis. Chronological age, however, is an imperfect surrogate. While it reflects population-level patterns, it fails to capture substantial inter-individual variation in the biological processes underlying aging (3). This limitation has motivated the development of ’biological age’ measures, which estimate an individual’s physiological state from clinical, molecular, or imaging data, independent of chronological age (4).

The rate of biological aging influences chronic disease risk, disability, and mortality. As populations age globally, the burden of age-related disorders, and in particular neurological disorders such as dementia and Parkinson’s disease, is expected to continue to rise (5). Quantifying biological aging may therefore enable the early identification of at-risk individuals and inform strategies to prevent disease and reduce disability (6). Previous studies using DNA methylation, proteomics, metabolomics, and brain magnetic resonance imaging (MRI) have shown the utility of biological age in predicting disease onset (7–10). However, most models were trained on a mix of healthy and non-healthy individuals, focused mainly on European ancestry, and relied on features such as MRI, proteins, methylation panels and metabolites, which are expensive to implement in the clinical setting (8–10).

To overcome the cost and accessibility barriers of omics data, researchers have developed age clocks based on clinical measurements such as blood counts and chemistry panels, which are extensively used in the clinic as routine indicators of general physical health (11), and have been shown to capture age-related processes and predict mortality and disease development (11–13). These models utilize common markers, such as aspartate aminotransferase (AST), alanine aminotransferase (ALT), or derived variables from hemograms, such as red blood cell-derived metrics (11,13). Yet, these cost-effective models have several limitations. For instance, PhenoAge leverages Cox proportional hazard models trained on mortality data (11), an approach that may prioritize markers of late-stage physiological dysregulation rather than the underlying molecular processes of aging. Meanwhile, BioAge uses the Klemera-Doubal method (KDM) that does not account for collinearity (correlation between features) (13). Finally, these two models have not been widely tested on non-European ancestries, so their generalizability remains unknown.

To address these gaps, we developed a novel biological age clock, with a particular emphasis on modeling neurological diseases, using a large, multi-ancestry cohort from the All of Us Research Program (N = 167,000) based on clinically measured blood and anthropometric biomarkers routinely measured in the clinic, offering an inexpensive approach to estimate biological age. We then assessed the association of the difference between this biomarker-estimated biological age (ClinAGE) and chronological age (Clinical-Biological Age Gap, ClinBAG) on aging-related traits spanning biological, sociodemographic, and cognitive domains. Next, we examined associations between ClinBAG and 434 neurological phenotypes and tested ancestry-specific heterogeneity across three major genetic groups: European, African, and Admixed American ancestry. Finally, we conducted a genome-wide association study (GWAS) to identify genetic variants influencing ClinBAG, revealing ancestry-specific effects shaped by population evolutionary history, and highlighted gene targets with potentially geroprotective effects.

## Results

### ClinAGE construction

**Figure 1** illustrates the study design. We used 22 clinical blood and anthropometric measurements from 14,328 sex-balanced individuals without neurological or psychiatric disorders to develop our ClinAGE model using LASSO and 5-fold cross-validation (Methods). All 22 features had non-zero coefficients (Figure 2d and Supplementary Table 1). Waist circumference had the largest positive coefficient (β=6.26), followed by systolic blood pressure (β=5.89), blood urea nitrogen (β=4.92) and mean corpuscular volume (MCV) (β=3.01). Weight (β=-5.47), diastolic blood pressure (β=-3.77) and blood protein levels (β=-2.41) had the highest inverse correlations with age. Following age-bias correction (Methods), ClinAGE was strongly correlated with age (Pearson R: 0.894, RMSE: 8.479) in the validation set (N = 2,866, Figure 2a). We then evaluated the performance of this model in a fully independent test cohort of healthy and non-healthy individuals from All of Us (N = 152,733) (Table 1). The model achieved good accuracy and generalizability (Pearson R: 0.858, RMSE: 8.983, Figure 2b and Supplementary Table 2). We then compared the performance of ClinAGE against PhenoAge, a widely used clinical age clock (Supplementary Figure 1). While the two were correlated (R = 0.772, N = 8,386), PhenoAge achieved a lower performance in All of Us (R: 0.696, RMSE: 16.5, MAE: 11.3). As C-reactive protein, a component of PhenoAge, was available in only 8,386 of 152,733 participants, precluding further evaluation of PhenoAge in downstream analyses.

**Figure 1.**
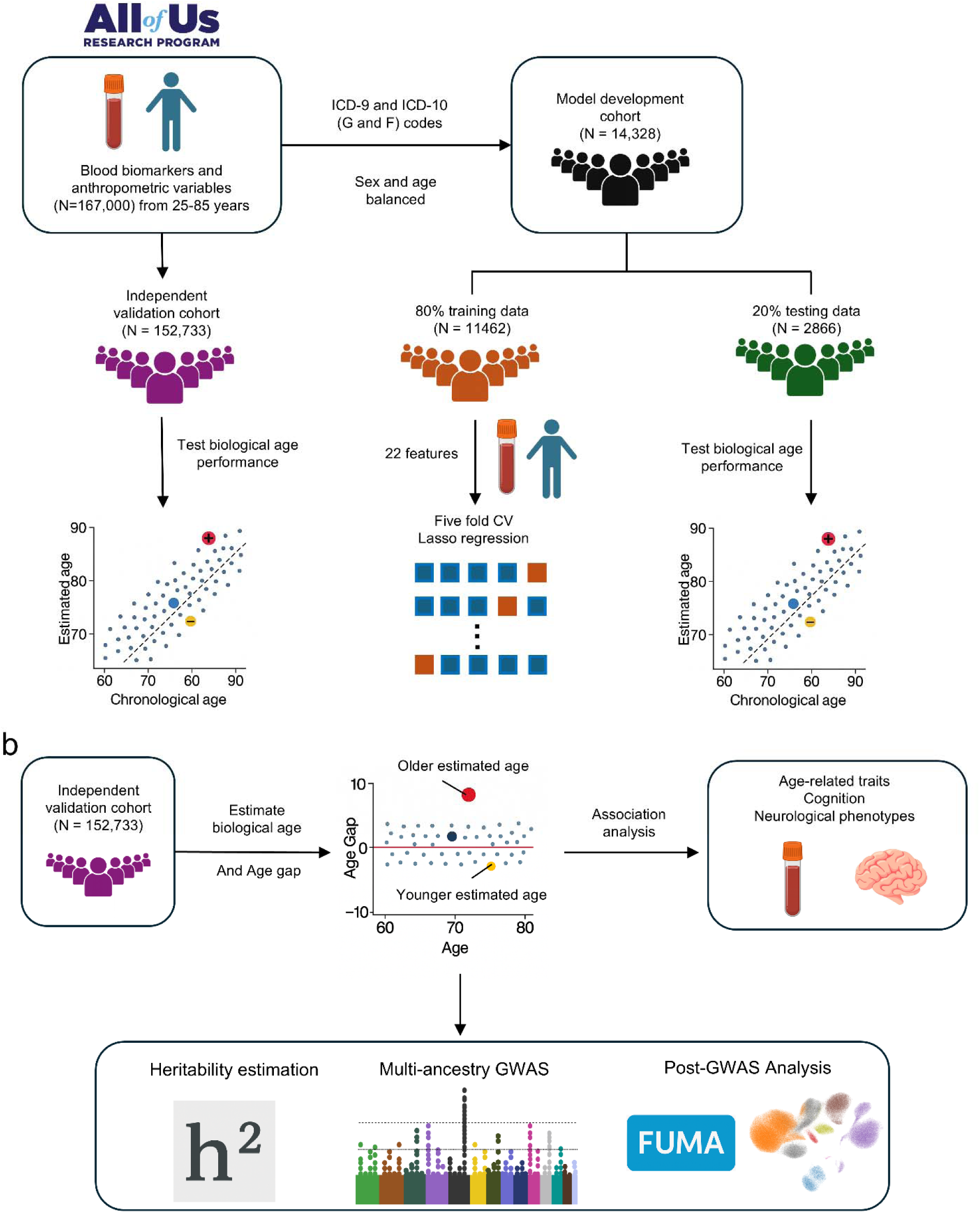
Overview of the study design and analytic approaches. a) All of US participants were split into a subset relatively healthy cohort with no neurological/mental phenotypes used for model development and a fully independent cohort that contains healthy and non-healthy individuals used for downstream analysis. The model development cohort was split into training and testing sets at an 80:20 ratio, age and sex balanced. In the training cohort, a Lasso linear regression model was trained to predict chronological age using 22 features. We evaluated the model performance in the test set, and we validated it using the fully independent cohort with 152,733 individuals. Biological age (ClinAGE) was calculated in the fully independent cohort. Biological age gap (ClinBAG) was calculated as the difference between ClinAGE and chronological age. We used linear and logistic regressions to test associations between ClinBAG and age-related biomarkers, cognition and neurological phenotypes. Finally, we conducted a GWAS on ClinBAG, estimated the heritability of this trait and performed downstream analysis using FUMA and single-cell RNA-seq datasets.

**Figure 2.**
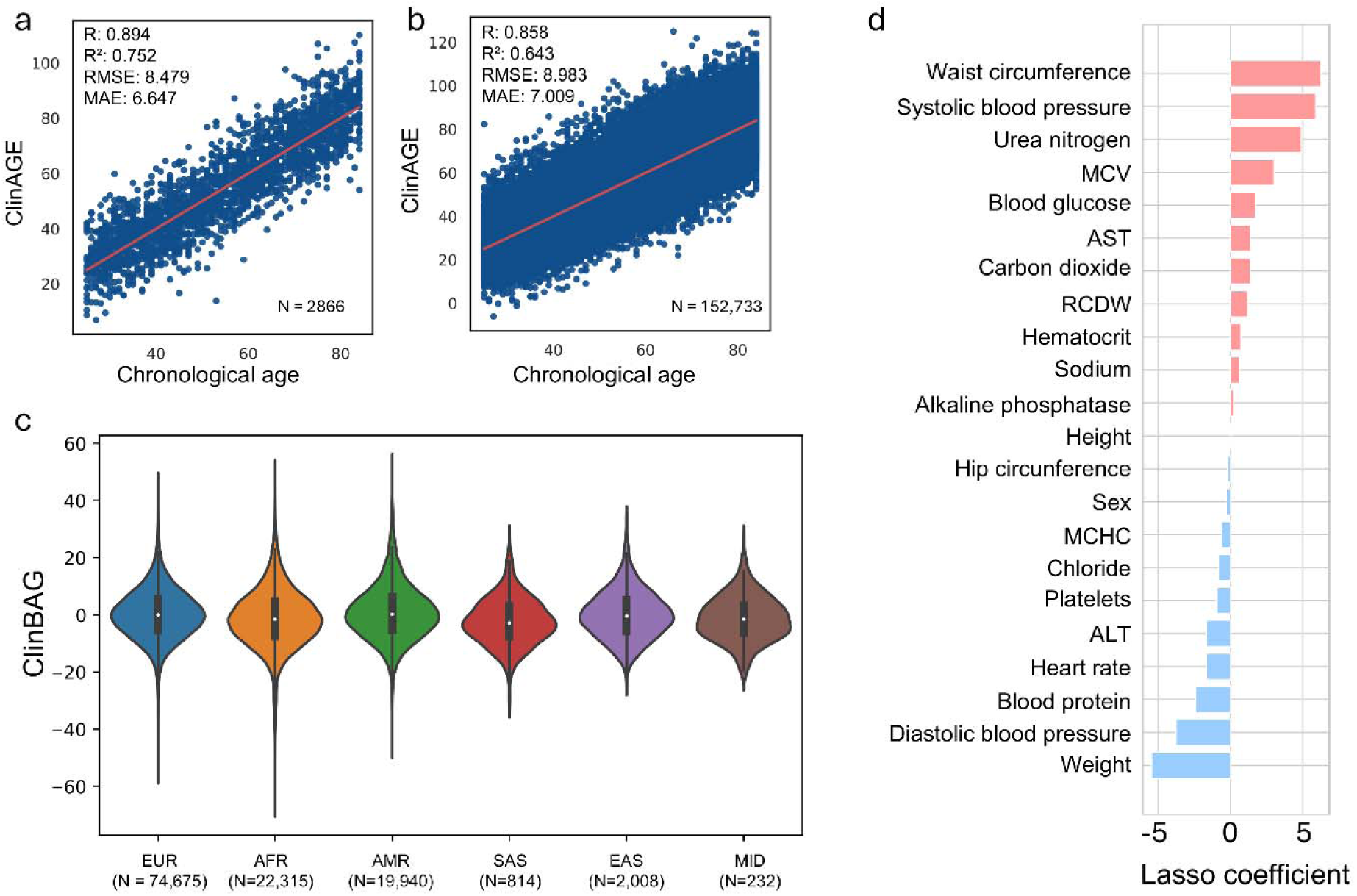
Model performance and features. a) Performance of the trained ClinAGE model in the testing set (N = 2866). b) Performance of the trained ClinAGE model in the fully independent cohort (N = 152,733). c) Distribution of ClinBAG according to genetic ancestry inferred using a random forest classifier: EUR (N = 74,675), AFR (N = 22,315), AMR (N = 19,940), SAS (N = 814), EAS (N = 2,008) and MID (N = 232). d) Lasso coefficients for every feature included in the ClinAGE model. Correlation coefficients shown in a-b are Pearson correlation coefficients. Violin plots in c, with center line, box limits and whiskers representing the median, interquartile range and minima/maxima within each group, respectively. RMSE: root mean squared error; MAE: mean absolute error. EUR: Europeans; AFR: Africans; AMR: Admixed Americans/Latinos; SAS: South Asians; EAS: East Asians; MID: Middle Easterners.

**Table 1.** Demographic characteristics of the population.

| Table 1. Demographic characteristics of the population |  |
| --- | --- |
|  | N = 152,733<sup>a</sup> |
| **Sex** |  |
| Female | 94,039 (61.57%) |
| Male | 58,694 (38.43%) |
| **Age** |  |
| mean (sd) | 55.63 (14.84) |
| Range (years) | 25 - 85 |
| **Genetic ancestry** |  |
| EUR | 74,675 (48.89%) |
| AFR | 22,315 (14.61%) |
| AMR | 19,940 (13.05%) |
| EAS | 2,008 (1.31%) |
| SAS | 814 (0.53%) |
| MID | 232 (0.15%) |
| Other/NA | 32,749 (21.46%) |
| **Neurological phenotype** |  |
| Yes | 134,196 (87.86%) |
| No | 18,537 (12.14%) |

<sup>a</sup>n(%); Mean (SD)

We calculated participants’ biological age gap (ClinBAG) as the difference between ClinAGE and chronological age in this test cohort. ClinBAG was not associated with chronological age and had similar distribution across five major ancestry groups: European ancestry (EUR), African ancestry (AFR), Admixed American ancestry (AMR), South Asian ancestry (SAS), East Asian ancestry (EAS) and Middle Eastern ancestry (MID) (Figure 2c).

### ClinBAG is associated with laboratory measurements, sociodemographic variables and cognitive decline

To validate ClinBAG, we evaluated its association with age-related biological, sociodemographic and cognitive function traits. Specifically, 1) 89 laboratory measurements that are not included in the construction of ClinBAG (e.g., estimated glomerular filtration rate, markers in urine, hemoglobin A1c, see Supplementary Table 3); 2) sociodemographic variables (income, education, health insurance); 3) self-reported physical health questions (difficulty concentrating, walking/climbing capacity and overall health and 4) cognitive function (emotion recognition, reaction time, concentration ability). Chronological age, sex and the first 10 genetic PCs were included as covariates. ClinBAG was associated with 35 out of 89 laboratory measurements at baseline (adjusted value < 0.05, Supplementary Table 3). Most baseline urine and blood marker levels increase with ClinBAG (Figure 3a). Estimated glomerular filtration rate (eGFR) showed the largest negative association with ClinBAG, consistent with the well-characterized age-related decline in renal function and indicating that ClinBAG captures variation in kidney aging beyond chronological age. To assess the predictive ability of ClinBAG, we repeated this analysis using the most recent follow-up measurement of each feature within a one-year window of the last follow-up date (median follow-up= 2.7 years, IQR=1.4 to 3.5 years). A total of 21 out of these 35 laboratory measurements showed consistent associations between the baseline and follow-up (Figure 3a and Supplementary Table 3). Interestingly, 5 laboratory measurements that were not associated with ClinBAG at baseline became significant in follow-up, including with increased levels of blood phosphate, anion gap, and decreased blood iron and total bilirubin, as well as urine specific gravity (Figure 3a and Supplementary Table 3).

**Figure 3.**
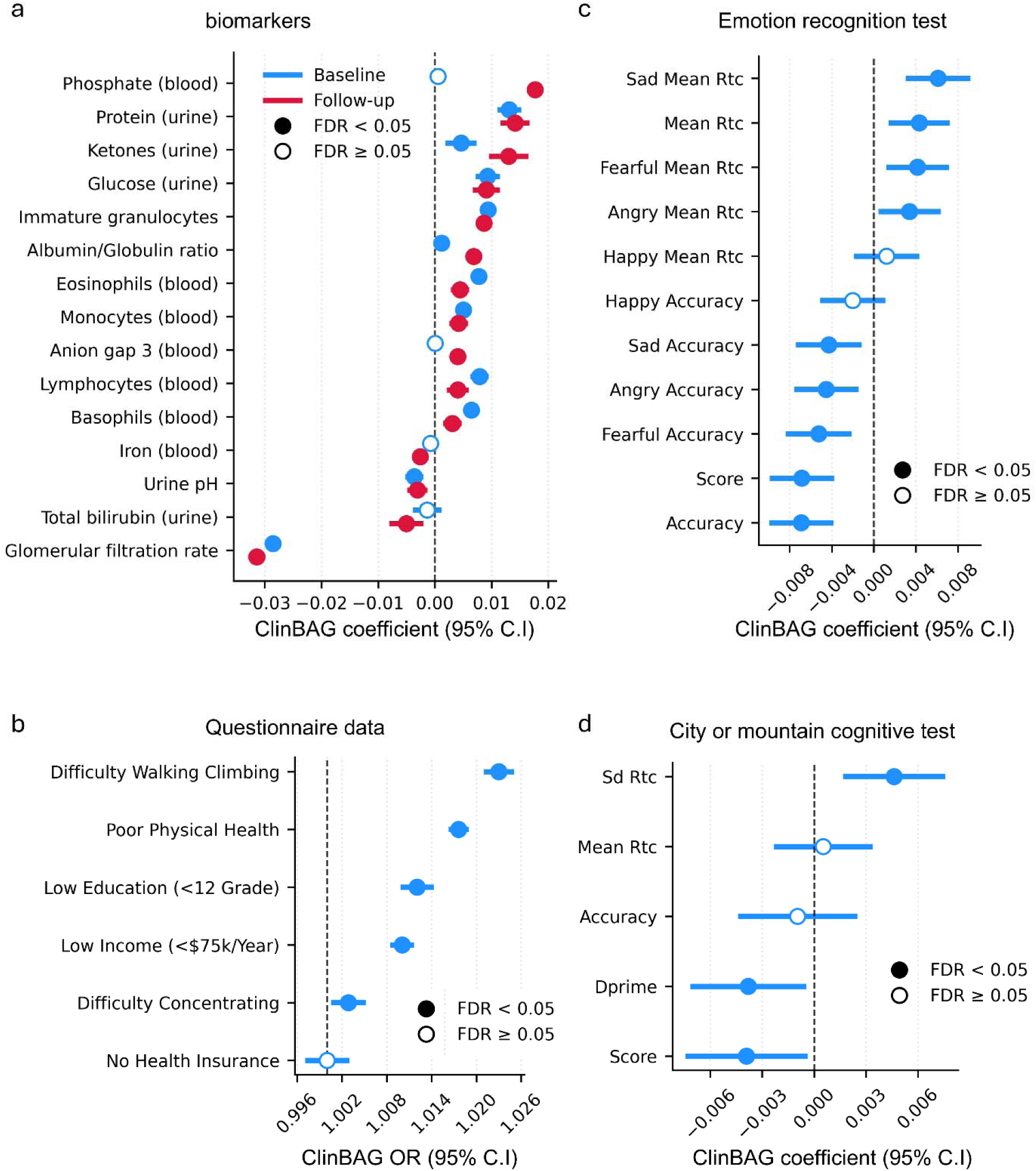
Association of ClinBAG with age-related biomarkers, sociodemographic variables and cognitive decline. a) associations between ClinBAG and age-related biomarkers in the fully independent cohort. Blue indicates the estimate at baseline and red at the last follow-up. b) Associations between ClinBAG and sociodemographic variables, including self-reported health, education and income. c) Associations between ClinBAG and the emotion recognition test by emotion and overall performance. d) Associations between ClinBAG and the City or Mountain cognitive test. All models used either linear or logistic regression and were adjusted by sex, chronological age and the first 10 genetic PCs. Rtc: reaction time; SD: standard deviation.

Higher ClinBAG was associated with increased difficulty walking/climbing, poor physical health, and difficulty concentrating (Figure 3b). Lower income (<75k USD per year) and education (<12 grade) were associated with higher ClinBAG, but not health insurance status. Individuals with higher ClinBAG also showed a reduced speed and accuracy of emotional recognition (Figure 3c), as well as lower attention and inhibition control (Figure 3d).

### Phenome-wide association analysis between BAG and neurological outcomes

We then tested the association of ClinBAG with 434 neurological and psychiatric phenotypes in a phenotype-wide analysis using Phecodes derived from ICD9 and ICD10 codes (Methods and Supplementary Table 4), adjusting for baseline chronological age, sex and 10 genetic PCs. In these case-control analyses, odds ratios are reported per 1-year increase in ClinBAG. P-values were adjusted using the false discovery rate (FDR) method. We identified 190 phenotypes significantly associated with ClinBAG (Supplementary Table 5). For most phenotypes, ClinBAG was associated with higher odds (Figure 4a and b). Notable exceptions included migraine (OR=0.993, 95% CI=0.9901-0.997, P-adjusted=0.0016), consistent with previous observations that the prevalence of migraine decreases with age (14). The phenotype with the strongest association with ClinBAG was metabolic encephalopathy, which is predominantly observed in older age groups (15) (OR=1.067, 95% CI=1.062-1.072, P-adjusted=5.92x10^-184^), followed by delirium (OR=1.058, 95% CI=1.052-1.065, P-adjusted=4.6x10^-63^) and obstructive sleep apnea syndrome (OR=1.017, 95% CI=1.015-1.019, P-adjusted=2.6x10^-49^). ClinBAG was also associated with neurodegenerative diseases, including dementia (OR=1.031, 95% CI=1.025-1.038, P-adjusted=2.13x10^-21^), Alzheimer’s disease (OR=1.019, 95% CI=1.008-1.031, P-adjusted=0.001) and Parkinson’s disease (OR=1.025, 95% CI=1.017-1.032, P-adjusted=3.7x10^-10^). We also observed a negative association of ClinBAG with schizophrenia (OR=0.992, CI=0.987-0.997, P-adjusted=0.019).

**Figure 4.**
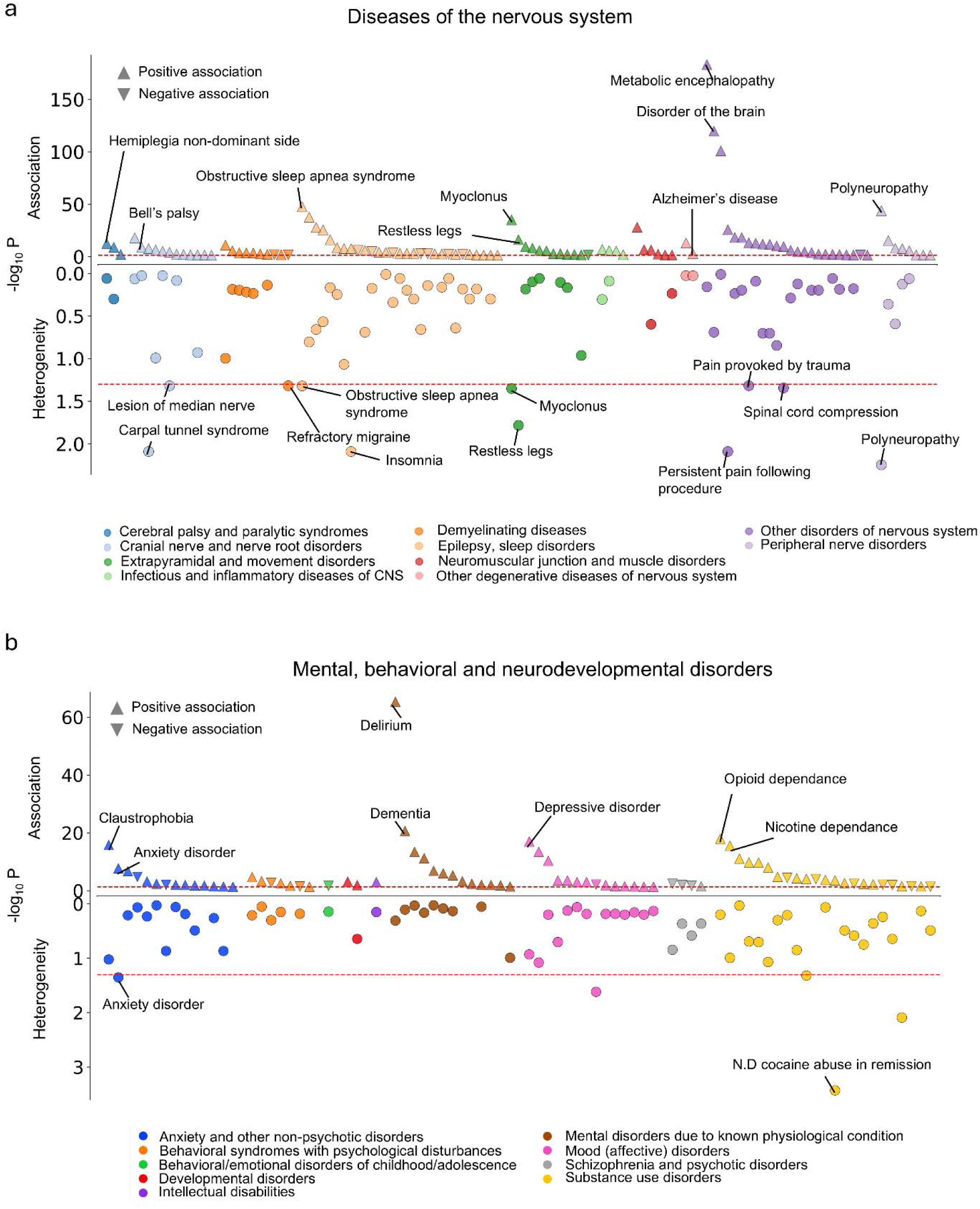
Association of ClinBAG with neurological and mental disorders. a) associations between ClinBAG and phenotypes of the nervous system (defined by ICD codes) tested by a logistic regression model adjusting for sex, chronological age and the first 10 genetic PCs. b) associations between ClinBAG and mental, behavioral and neurodevelopmental phenotypes assessed with a logistic regression adjusted by sex, chronological age and the first 10 genetic PCs. For a and b, adjusted P-values are shown for the association test (upward) and Heterogeneity (downward) between EUR, AFR and AMR, assessed by a Cochran’s Q test.

For ClinBAG-associated phenotypes, we next performed an ancestry-specific analyses and evaluated heterogeneity. We detected significant heterogeneity for 16 phenotypes (Adjusted P-value <0.05, Figure 4b, 5c, Supplementary Figure 2 and Supplementary Table 6), whereby, BAG showed a significant positive association with insomnia, anxiety and restless legs in EUR and AMR, but not significant in AFR (despite similar sample sizes for AMR and AFR, Supplementary Figure 2). Moreover, we found a negative association of ClinBAG and episodic mood disorder only in AMR (Figure 4 and Supplementary Figure 2).

To reduce the risk of reverse causation, we re-examined the ClinBAG-associated phenotypes using time-to-event analyses restricted to incident cases arising at least one year after recruitment and ClinBAG-component measurement (Methods, Figure 5a). The median follow-up was 2.7 years (IQR=1.4 – 3.5 years). Each 1-year increase in ClinBAG was associated with an increased hazard of delirium (HR=1.053, 95% CI=1.044-1.063, P-adjusted=1.08x10^-28^), tremor (HR=1.018, 95% CI=1.009-1.027, P-adjusted=0.00024), dementia (HR=1.020, 95% CI=1.011-1.029, P-adjusted=0.000016), Parkinson’s disease (HR=1.014, 95% CI=1.002-1.027, P-adjusted=0.023), among others (Figure5a). Similarly, high ClinBAG was associated with a decreased risk of developing migraine (HR=0.991, CI=0.985-0.996, P-adjusted=0.00087) and schizophrenia (HR=0.987, CI=0.974-0.999, P-adjusted=0.044, Figure 5a). The cross-sectional association between ClinBAG and post-traumatic stress disorder was not replicated in time-to-event analysis, although the number of incident cases was low (n = 1639). This suggests the direction of effect may be from post-traumatic stress disorder to accelerated biological aging, potentially mediated by associated factors such as substance use and comorbidities. Kaplan Meier curves showed a clear separation and increased incidence of dementia in individuals with a higher ClinBAG, and the opposite trend for schizophrenia (Figure 5b).

**Figure 5.**
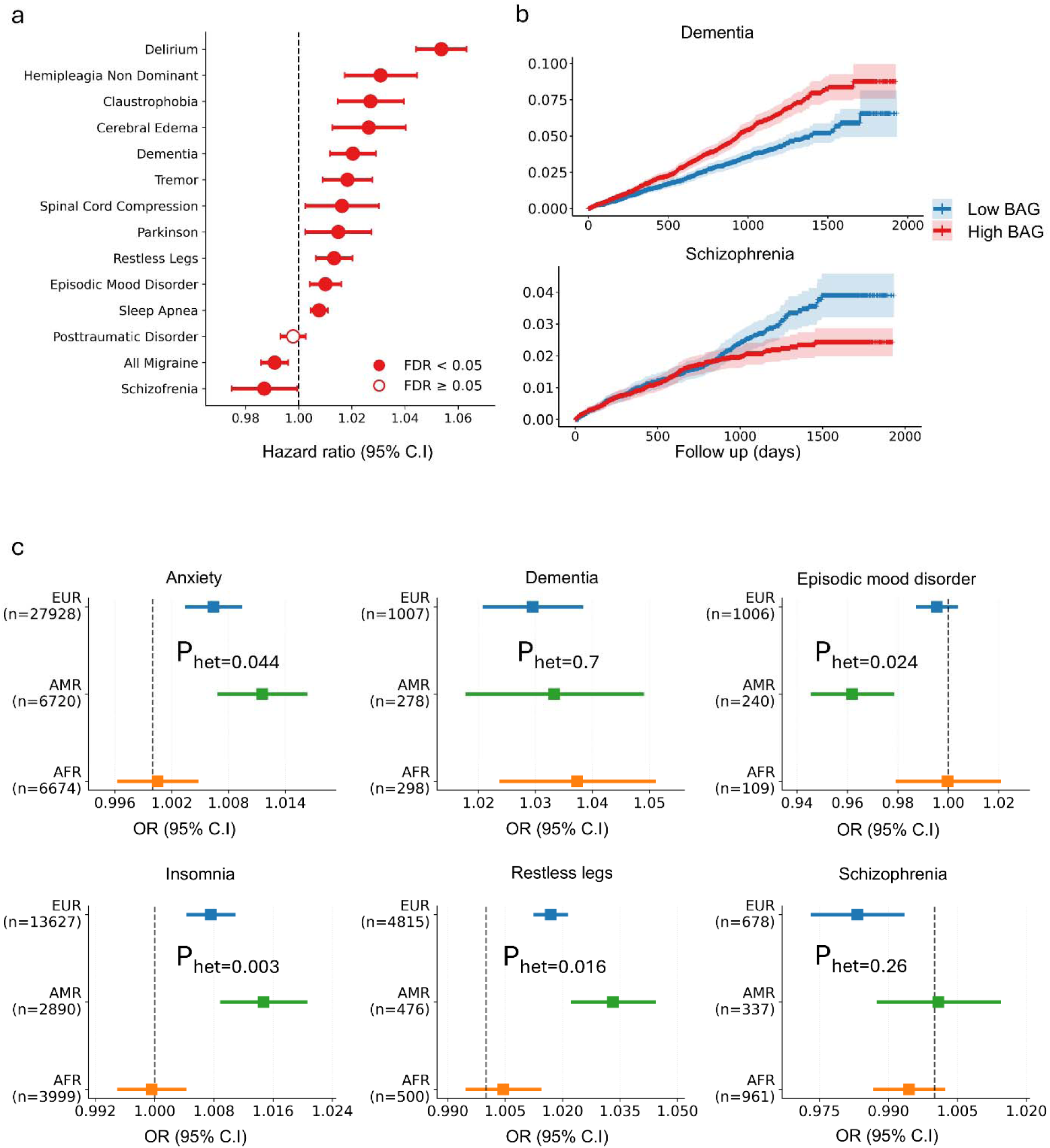
ClinBAG increases the risk of developing neurological phenotypes. a) Associations between ClinBAG and neurological phenotypes using Cox proportional hazard models, adjusted by sex, chronological age and the first 10 genetic PCs. b) a cumulative incidence plot where individuals were divided by the median ClinBAG . All plots show cumulative incidence density of events at a given timepoint on the Kaplan-Meier survival function, with 95% confidence intervals shown in lighter shading. c) Comparison of effect sizes from the logistic regression stratified by ancestry for EUR, AFR and AMR. P-values were computed using a Cochran’s Q test and corrected using the FDR method.

### Multi-ancestry GWAS of ClinBAG

To map the genetic architecture of ClinBAG, we conducted GWAS for ClinBAG in three populations: EUR, AFR and AMR, each with at least ∼20,000 individuals. Covariates were sex, the first 10 genetic PCs and sequencing site. We observed non-zero significant heritability for ClinBAG in AFR (20.7%), EUR (20.9%) and AMR (6%, likely underpowered). We found two genome-wide significant loci (p<5x10^-8^) in AFR, five in EUR and four suggestive loci in AMR (p<1x10^-7^) populations (Figure 6a, b and c). We did not find evidence suggesting residual population stratification or cryptic relatedness via the linkage disequilibrium (LD) score regression intercept (1.0143 in AFR, 1.0143 in EUR and 0.980 in AMR). Genomic control factor (lambda GC) was 1.025 for AFR, 1.074 for EUR and 1.008 for AMR, suggesting no inflation.

**Figure 6.**
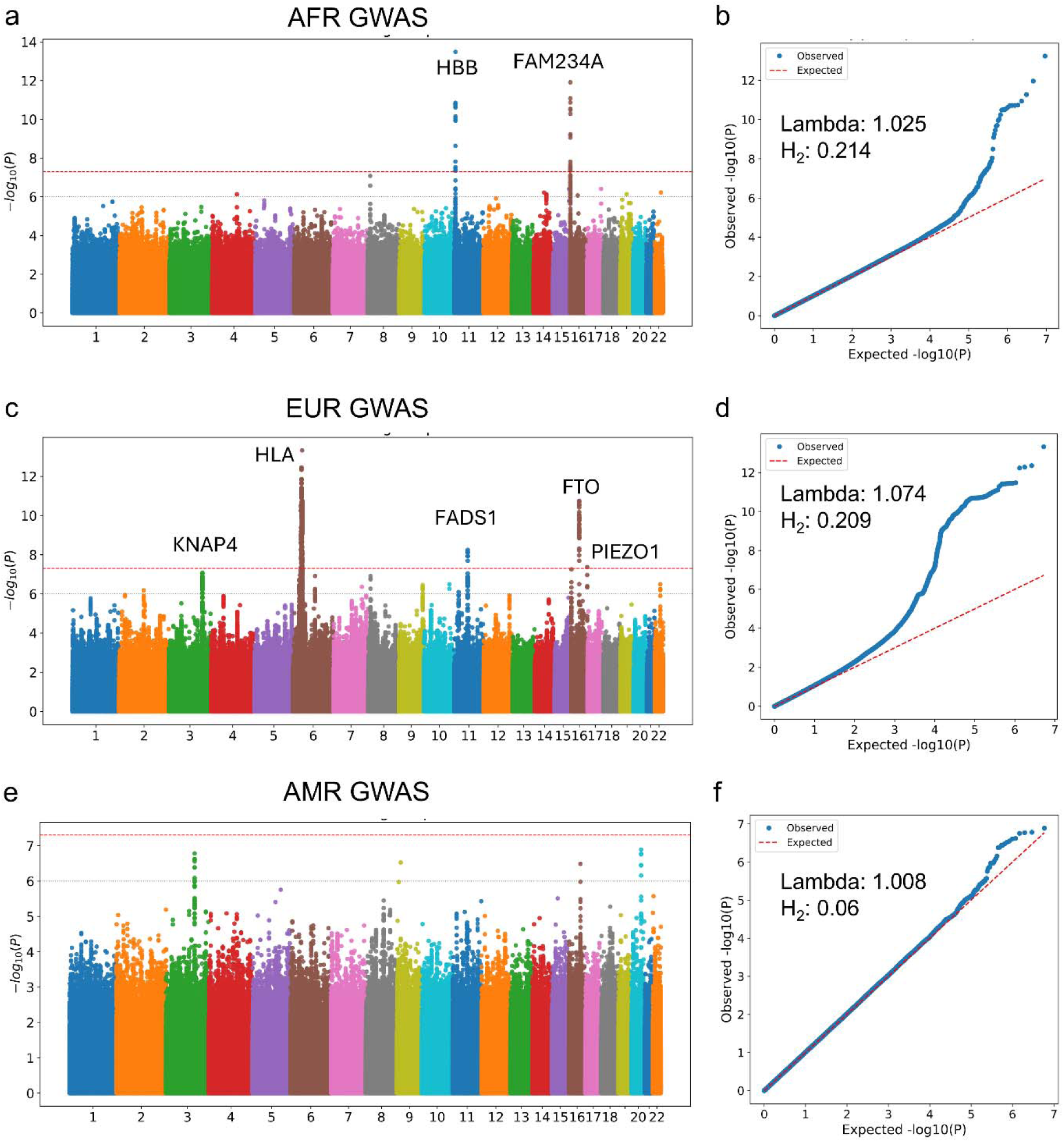
Multi-ancestry GWAS on ClinBAG. a) Manhattan plot of the African GWAS on ClinBAG, highlighting two hits in *HBB* and *FAM234A*. b) Q-Q plot, lambda GC and heritability estimate for Africans. c) European GWAS on ClinBAG, showing significant associations with *KNAP4, HLA, FADS1, FTO* and *PIEZO1*. d) Q-Q plot, lambda GC and heritability estimate for Europeans. e) Admixed Americans/Latinos GWAS on ClinBAG with 4 nominally significant associations. f) Q-Q plot, lambda GC and heritability estimate for admixed Americans/Latinos. The dashed red line denotes the genome-wide significant threshold (5x10^-8^). The grey line denotes the nominally significant threshold (5x10^-7^).

### GWAS of ClinBAG in AFR

In the AFR cohort, the most significant signal was identified on chromosome 11 (chr 11). The lead variant, rs334, is an exonic single nucleotide polymorphism (SNP) with a CADD score of 13.85. Notably, this variant is the causal mutation for sickle cell anemia and maintains a minor allele frequency (MAF) >1% exclusively in AFR in the All of us cohort. It has previously been associated with various blood-related phenotypes in the GWAS Catalog such as red blood cell distribution width (RDW), MCV, mean corpuscular hemoglobin concentration (MCHC), among others (Figure 6a; Supplementary Table 7).

On chr16, we identified a locus near the gene FAM234A. Nearby genes—including *NPRL3, HBA1, HBA2*, and *HBZ*—are primarily involved in erythroid cell function, suggesting this locus may regulate genes critical for hemoglobin production. The lead SNP rs13331259 (*FAM234A*) has been associated with traits used to train our biological age model, such as RDW, platelet volume, and iron deficiency anemia in the GWAS Catalog (Supplementary Table 8). Furthermore, this variant acts as an GTEx eQTL for *AXIN1* (P-adjusted=0.002), a protein involved in the Wnt–NAD+ pathway—a well-established mechanism in biological aging.

To identify potential target genes for non-coding variants within the chr16 locus, we performed fine-mapping using SuSiE. The 95% credible set contained five variants in two genes (*FAM234A* and *HBA1*), with rs13331259 (*FAM234A*) emerging as the most plausible candidate (posterior inclusion probability = 0.73, Supplementary Table 8). Subsequent gene-set aggregation using the MAGMA SNP2GENE function identified four significantly associated genes: *ITFG3, NPRL3, LUC7L,* and *ARHGDIG*. Collectively, these prioritized genes belong to a genomic cluster involved in hemoglobin production.

### GWAS of ClinBAG in EUR

In the EUR cohort, the most significant GWAS signal mapped to the HFE locus on chr6. Additional signals were identified on chromosomes 3, 11, and 16 (Supplementary Table 9). None of the loci were found significant in the AFR GWAS.

Three of the five identified genomic risk loci had been previously reported in European GWAS for biological age clocks trained on blood biomarkers (e.g., PhenoAge): the FADS1/FADS2 locus (chr 11), the HLA locus (chr 6), and the FTO locus (chr 16) (16). In each case, our lead variants were in high linkage disequilibrium (LD) with previously reported signals, validating our European-centric ClinBAG. Furthermore, all lead variants have known associations with at least one biomarker used as features for ClinBAG (Supplementary Table 10).

For the novel chr3 locus, we prioritized rs4679654 due to its high CADD score (21.6) and being a cis-eQTL for *KPNA4.* Among 13 genes located in this locus, rs4679654 also influences the expression of *SMC4 (*P-adjusted= *5.3x10^-5^), TRIM59 (*P-adjusted= *4x10^-6^),* and *B3GALNT1* (P-adjusted=6.2x10^-8^). On chr11, we highlight rs174538 (CADD = 14.1), a cis-eQTL for *FADS1* (P-adjusted=1.6x10^-6^), which encodes a key protein in lipid metabolism. Finally, on chr16, we prioritized two variants across distinct risk loci: rs7202296 (CADD = 18.01), a cis-eQTL for *FTO* (P-adjusted=7.8x10^-13^), and rs8052370 (CADD = 7.0), which acts as an eQTL for both *PIEZO1* (P-adjusted=0.002) and *APRT* (P-adjusted=0.013).

Moreover, the MAGMA gene-based test found 13 genes that were significantly enriched for ClinBAG in the European population (P-value < 5x10^-6^, Supplementary Table 11), including *HLA-DRA, FADS1-2, NPRL3, FTO*, and *MIEF1*. This gene-based association test also revealed *NPRL3* to be enriched for both EUR and AFR ClinBAG GWAS, where individual SNPs in *NPRL3* were only nominally significant in the EUR GWAS.

Additionally, we found 4 nominally significant independent risk loci for AMR in chromosomes 3, 9, 16 and 20 (P-value <1x10^-7^). Interestingly, the chr3 and chr16 loci were independent from the EUR and AFR GWAS. Lead variant rs9822469 in chr3 is located in *RYK*, a key gene for hematopoietic stem cell differentiation. Lead variant rs75614553 in chr9 is located at the pseudogene *RP11-54D18.3*. We prioritized the variant rs12597011 in chr16 as an eQTL for *CALB2* (P-adjusted=0.045), a key gene related to obesity.

### Gene set enrichment analysis and effector cell types

In the EUR ClinBAG GWAS, we identified 108 genes, and enrichment of these genes was strongest in the pancreas, spleen, stomach, thyroid and blood vessels (Figure 7a, Supplementary Table 12). Enrichment analysis linked these genes to cholesterol metabolism pathways, lipid storage, acetyl-CoA metabolic process, and negative regulation of cell growth (Figure 7b and Supplementary Table 13). We then evaluated the enrichment of the EUR gene-set in a single-cell RNA-seq dataset from peripheral mononuclear cells (PBMCs) from healthy individuals (17) with ages spanning 25 to 85 years (Figure 7c). Interestingly, the expression of these prioritized gene increased with age in double negative (DN) T cells (Figure 7d). These genes were also significantly enriched in hematopoietic progenitor cells, independent of age (Figure 7e).

**Figure 7.**
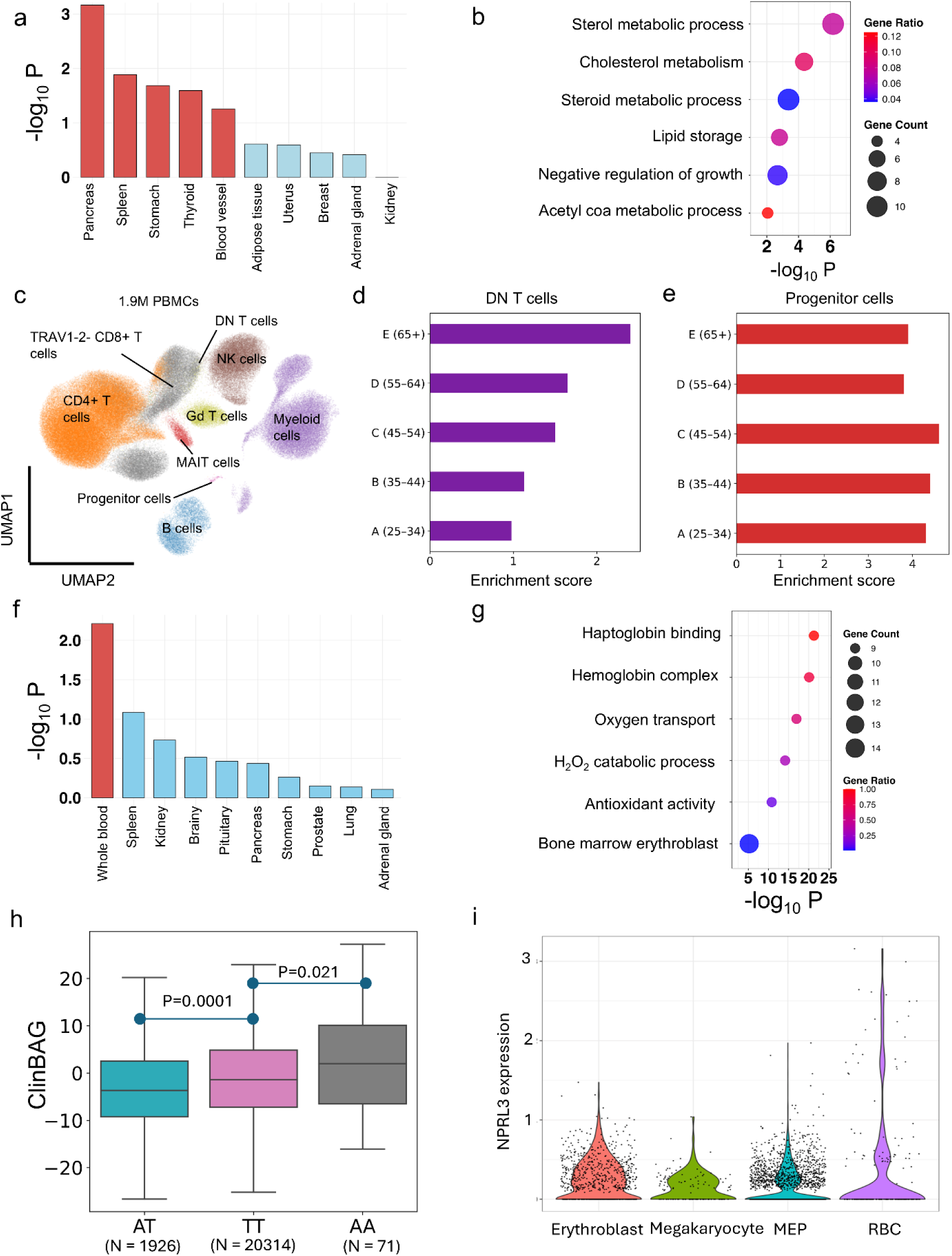
Post-GWAS downstream analysis using FUMA and scDRS. a) Enrichment of expression of 13 genes prioritized by MAGMA from the European ClinBAG GWAS in 54 GTeX tissues. Red indicates tissues significantly enriched after FDR correction. b) enrichment analysis in Gene Ontology pathways for the European GWAS, highlighting sterol metabolic processes and lipid storage. The X-axis displays the adjusted p-value for every pathway. Dot size shows the number of genes from that pathway present in the evaluated gene-set, and gene ratio is the division of gene count by the total number of genes in that pathway. c) UMAP representation of a single cell dataset with 1.9M cells from PBMCs in healthy Europeans from 25-65+ years. d) enrichment of the European MAGMA gene-set in DN T cells in an age-dependent manner. e) enrichment of the European MAGMA gene-set in progenitor cells regardless of age. f) Enrichment of expression of 13 genes prioritized by MAGMA from the Africans ClinBAG GWAS in 54 GTeX tissues, highlighting blood as the only significant tissue. g) enrichment analysis in Gene Ontology pathways, similar to b, highlighting the hemoglobin complex, oxygen transport, and erythroblasts from bone marrow. h) ClinBAG distribution across different genotypes for the sickle cell disease variant rs334. TT (healthy), AT (sickle cell trait) and AA (sickle cell disease). P-values were computed using a linear regression, with TT as reference and adjusting by sex, chronological age and the first 10 genetic PCs. i) Violin plot showing the distribution of *NPRL3* expression in a single-cell dataset of bone marrow and PBMCs from healthy European individuals. DN T cells: double negative T cells; MEP: Megakaryocyte–erythroid progenitor; RBC: red blood cell.

In the AFR ClinBAG GWAS, we identified 42 genes, and they were significantly enriched in whole blood (Figure 7f). Enrichment analysis for this AFR gene set revealed pathways associated with oxygen transport, hemoglobin complex, antioxidant activity and erythroblasts from bone marrow (Figure 7g). The difference between EUR and AFR gene-set enrichment analyses showed ancestral diversity in biological aging, reflected in same features used in constructing ClinBAG that may weigh differently across ancestry groups.

### HBB and NPRL3

We further investigated two loci from ClinBAG GWAS: the AFR specific *HBB* locus and *NPRL3*, which was shared across AFR and EUR. At the *HBB* locus, we compared ClinBAG across genotypes of the sickle cell variant rs334: TT (reference), AT (sickle cell trait or carriers) and AA (sickle cell disease). Homozygous individuals (AA) had higher ClinBAG than non-carriers (β=2.76, 95% CI 0.41–5.10, P=0.021, Figure 7h), consistent with the known effects of sickle cell disease on blood biomarkers and accelerated aging (18–21). Unexpectedly, heterozygous carriers (AT) had lower ClinBAG than non-carriers (β=−2.19, 95% CI −2.66 to −1.72, P=1×10⁻□). To understand this, we compared individual biomarker levels across genotypes and found that carriers and non-carriers had similar profiles, except for mean corpuscular volume (MCV), which was lower in heterozygous carriers (β=−0.54, 95% CI −0.60 to −0.49, P=6.6×10⁻□², Supplementary Figure 3). Given the positive coefficient of MCV in the ClinBAG model, this likely explains the lower biological age estimate in carriers.

Given the importance of NPRL3 for erythropoiesis (22), we investigated the expression of *NPRL3* in a single-cell RNA-seq dataset of bone marrow from healthy individuals of EUR ancestry (23). We found a high expression of *NPRL3* in erythroblasts, megakaryocytes, erythroid megakaryocyte progenitor and red blood cells (Figure 7i and Supplementary Figure 4).

## Discussion

We developed ClinBAG, a multi-ancestry biological aging clock derived from routine clinical blood biomarkers and anthropometric measures in the All of Us cohort, and demonstrated its accuracy and generalizability across multiple ancestries (EUR, AFR, and AMR). Through comprehensive cross-sectional and longitudinal analyses, we demonstrate that ClinBAG is associated with variation in key general health, cognitive function, and sociodemographic factors, and predicts future change in clinically-relevant biomarkers such as eGFR. Our study also provides evidence that ClinBAG is a risk factor for several neurological diseases and age-related functional status. Finally, ClinBAG showed significant heritability with distinct genetic architectures across EUR, AFR, and AMR ancestries. Similarly, gene-level analyses revealed ancestry-specific genes and processes that contribute to biological aging.

In contrast to previous models restricted to mid-to-late life, such as those trained on UK Biobank data (40-70 years) (24,25), our ClinBAG includes individuals with a wider age range (25-85 years) resulting in improved generalizability and applicability. We also found a better performance (r=0.85) of our ClinBAG than previous clocks constructed from clinical routine clinical measurements such as PhenoAge (r=0.696) (11). These models are trained on mortality data, capturing the late-stage biological consequences of aging rather than molecular processes that influence risk. In addition, they were trained on on-healthy individuals, limiting their interpretation. In contrast, our approach offers a simple and low-cost alternative to capture meaningful accelerating aging processes that only rely on 22 routine clinical measurements and chronological age for their estimation, trained on relatively healthy participants, offering simplicity and feasibility in a clinical setting.

When comparing our findings with another multi-ancestry biological aging study using the All of Us cohort, which used telomere length as an aging biomarker, most of the features in our ClinBAG model, such as blood pressure, height, weight, glucose, and heart rate, are associated with telomere length, highlighting their orthogonal contribution to aging (26). Most importantly, our ClinBAG model was applied to disease phenotypes and provided previously unrecognized evidence of heterogeneous effects of biological clock between ancestries (Supplementary Figure 2).

Both cross-sectional and longitudinal analyses confirmed a negative association between ClinBAG and migraine. We interpret this to reflect the influence of biological aging on migraine susceptibility, consistent with and independent of the well-established decline in migraine prevalence with chronological age. Consistent with this, prior studies showed no association between migraine and accelerated epigenetic aging (27) or cognitive decline (28). Reports of older brain age in migraine patients using MRI (29,30) may reflect organ- and/or modality-specific effects rather than systemic biological aging. Similarly, ClinBAG was negatively associated with schizophrenia (cross-sectional and longitudinal evidence). In agreement, schizophrenia incidence decreases with age, specifically with a lower rate after the fifth decade (31). A previous report of older epigenetic age in schizophrenia patients may reflect a modality specific effect rather than systemic biological aging (32). Another study found a higher PhenoAge and BioAge in patients with Schizophrenia in the UK biobank (33). In addition, they reported an effect modification mediated by body mass index, a variable that we used indirectly in our ClinBAG (weight and height). Our cross sectional results are in concordance with previous observations that antipsychotic medications increase heart rate (34) and appetite (35), and they could alter some of the features we used to train our model (heart rate, weight, systolic blood pressure, Supplementary Figure 5). Additional evidence suggests that some of these observations, including increased heart rate (36), fasting glucose (37), BMI (38) and chronic inflammation (39), may present in the prodromal phase prior to diagnosis and contribute to our findings in the longitudinal analysis.

For the AFR GWAS, the Rs334 variant was associated with ClinBAG. Importantly, individuals with sickle cell disease (Rs334 AA genotype) showed a higher ClinBAG (Figure 7h), consistent with previous studies showing that sickle cell anemia accelerates aging (18–21), even classifying sickle cell anemia as an aging syndrome (40). However, sickle cell trait individuals (rs334 AT genotype) showed a decreased ClinBAG. Sickle cell trait individuals have lower MCV, hematocrit, RDW and HbA1c levels (41). Given the positive Lasso coefficients for MCV, blood glucose, carbon dioxide and hematocrit in our age clock, it is expected that carriers of rs334 would have a lower ClinBAG. Moreover, a study on telomere length in the UK Biobank found an association between the rs334-A allele and longer telomeres, but they concluded that this SNP alters the quantitative polymerase chain reaction (qPCR) assay that measures the normalizing control (42). Whether rs334 heterozygosity is associated with longevity or not remains inconclusive.

Previous GWAS of age clocks trained on routine clinical biomarkers did not observe the distinct effect of rs334 because it is absent from European populations (11,13,16). The unique selective pressures related to Malaria resistance in African populations led to the rise of the rs334 variant frequency, which is also associated with red blood cell metrics, key features of ClinBAG. This highlights the importance of incorporating multi-ancestry cohorts in age clock validation.

EUR ClinBAG GWAS revealed a signal in the HFE locus, near the HLA region, suggesting that we captured certain immune-related aging processes, consistent with our positive associations between ClinBAG and immature granulocytes, eosinophils, monocytes, basophils and lymphocytes (Figure 3A). Our single-cell enrichment analysis found a strong correlation between the prioritized genes and their expression in DN T cells in older adults (65+), and an increased expression of this gene set across age (Figure 7d). DN T cells have been implicated in several age-related conditions, such as Alzheimer’s disease and Parkinson’s disease (44,45). Specifically, DN T cells increase with age in mice models of Alzheimer’s disease, including broad DN T cells (CD4- and CD8-) (46) and specific subpopulations (effector/effector memory and KLRG1+ DN T cells) (44). DN T cells have also been associated with autoimmune diseases due to their role in regulating alloreactive, xenoreactive and autoreactive T cells (47); as well as promoting neuroinflammation (48). Interestingly, a few studies have found increased DN T cells in peripheral blood following a surgery or stroke event, and they can pass the blood-brain barrier (BBB) to amplify neuroinflammation produced by microglia releasing TNF-alpha (49). Higher proportions of DN T cells have been found in the peripheral blood of older adults (50), consistent with our observation. DN T cells decrease cytotoxic effect in the endocervix (51) and increase antiviral response and innate immune system, promoting inflammation (52).

We highlight *NPRL3* as the only gene consistently associated with biological aging across ancestries. Germline mutations in *NPRL3* are associated with epilepsy and malformations of cortical development (53). *NPRL3* is an inhibitor of the mTORC1 pathway, which promotes cell proliferation by increasing global protein synthesis and other anabolic processes, thought to cause irreversible cell senescence, oxidative and proteostatic stress and therefore aging (54–56). *NPRL3* is expressed in hematopoietic progenitors, and its expression is further boosted in erythroid lineages by alpha-globulin enhancers (22). *NPRL3* introns contain 3 out of 4 alpha-globulin enhancers for humans. Our single-cell analysis demonstrated *NPRL3* expression in erythroid progenitors and mature red blood cells. *NPRL3* controls autophagic flux (critical in erythroblasts) and regulates mTORC1 response to amino acid and iron availability, establishing a critical role in erythropoiesis (22). Its role in erythropoiesis could explain why *NPRL3* is a hit for both EUR and AFR, given that we used red-blood cell-related features in our aging model. mTOR pathway has been pharmacologically targeted by rapamycin to prevent aging (57) and phase1 clinical trials on rapamycin either for aging or Alzheimer’s disease have shown decreased red blood cell biomarkers such as MCV, RDW and MCHC (used as features in our aging model) (58,59), highlighting a role of erythropoiesis on aging and the utility of using red blood cell-related biomarkers for biological age estimation (60).

We acknowledge several limitations of our study: as we only focused on neurological and behavioral diseases, our ClinBAG may not be the best model for evaluating other aging related diseases. In the feature selection process, we applied a linear model with L1 regularization, which does not capture non-linear relationships between our features and aging. Moreover, most GWAS hits are associated with features used to train our model, which might reflect associations with features in our model and not only aging. Finally, due to sample size limitations, we could not include other major populations, such as East Asians, in the evaluation and genetic mapping of ClinBAG.

In summary, we demonstrated that a cost-effective biological age clock trained on blood and anthropometric measurements is associated with cognitive decline and tracking age-related biomarkers. We confirmed the association and predictability of neurological and mental diseases, while highlighting ancestry heterogeneity. Our genomic mapping uncovered genes important for biological aging, such as *NPRL3* and globulin-related genes and their possible molecular mechanisms, which may point to promising targets for geroprotective therapies.

## Methods

### All of Us Cohort

The All of Us Research Program (AoU) is a diverse cohort of individuals from different backgrounds, including Europeans, Africans, admixed Americans (Latin Americans) and Asians (South and East Asians) (61). Briefly, adults aged 18 years or older who consented and resided in the U.S were eligible. All participants consented to be part of the study and provided access to their electronic health record (EHR). Access to All of Us was granted through McGill University (IRB: 2107073).

### Blood biomarkers

Information about blood biomarkers, including lipid levels, general immune populations, metabolites, etc. (Supplementary Table 1), was retrieved from the “All of Us” workbench. These blood biomarkers were extracted from electronic health records (EHR), and most individuals contain multiple measurements. To align the time of measurement as closely as possible with the recruitment date (baseline), we selected only biomarkers that were measured within one year of the recruitment date for each participant (either one year before or after). If a blood biomarker was measured outside of this time, it was set to missing in the dataset. This dataset included 301,000 participants and 56 variables.

### Anthropometric measurements

We retrieved measurements regarding height, weight, systolic and diastolic blood pressure, heart rate, waist circumference, and hip circumference that were directly measured by the All of Us team at baseline. We selected the average value across successive measurements for robustness.

### Biological age estimation

We combined routine clinical markers (n=56) with anthropometric measurements (n=7) from 301,000 individuals. We excluded features with more than 50% of missing data, resulting in a dataset with 22 features (14 clinical routine markers, 7 anthropometric measurements and sex). We then removed individuals with more than 35% of missing data, resulting in 179,313 individuals. After, we utilized ICD-9 codes ranging from 290-359 and ICD-10 codes (F* and G*) to exclude individuals with neurological and psychiatric conditions, as recommended elsewhere (62). A total of 36,060 healthy individuals were retained for downstream analysis. To ensure a balanced training dataset across ages, we defined the following age groups: 26-35, 36-45, 46-55, 56-65, 66-75, 76-85. Individuals with ages <26 and > 86 were excluded due to low sample size. We then selected a subsample from this cohort of 36,060 healthy individuals, ensuring the same number of individuals in each age group, and an equal number of females and males. The final healthy cohort included 14,328 individuals (with 1194 males and 1194 females per age group). We split this dataset into 80% for training (N = 11,462) and 20% for testing (N = 2,866) in a sex and age-balanced way. In the training cohort, we first applied inverse rank normalization to obtain features with comparable magnitudes. To deal with missing information, we applied a K-nearest-neighbour (KNN) imputation method with K=5, as implemented in the KNNImputer function (with K=5) in sklearn. This trained KNN imputer was used to impute missing data in both the testing and the independent cohort.

We then applied a Lasso linear regression (L1 regularization) to perform feature selection and evaluate the performance of the model in predicting chronological age. We used a 5-fold cross-validation to find the best alpha hyperparameter for the Lasso regression with the LassoCV function in sklearn. We then used this hyperparameter to estimate the Lasso coefficients in the full training dataset using the Lasso.fit() sklearn function.

### Age bias correction

Biological age estimation possesses a bias where it overestimates biological age in younger individuals and underestimates it in older individuals. This generates a negative correlation between biological age gap (ClinBAG) – the difference between biological age and chronological age – and chronological age. To remove this bias, we regressed the biological age on chronological age and used the intercept (α) and the slope coefficient (β) of this regression to apply a statistical correction on the biological age in the training dataset as described in more detail elsewhere (9), using the following equation:

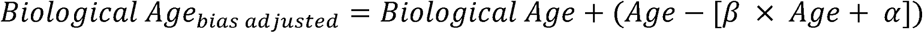

The KNN imputer and age bias correction were fitted on the training dataset and used to impute and correct on the testing and independent datasets to prevent data leakage.

#### Model performance

Performance in the training dataset was computed as the average performance across five folds after age-bias correction. We calculated Pearson correlation (R), correlation coefficient (R^2^), mean absolute error (MAE), and root mean square error (RMSE) between our estimated age and chronological age as performance metrics. After estimating the corrected biological age using the Lasso coefficients from the training dataset, we computed the same metrics in the testing dataset. In addition, we performed independent validation using the remaining cohort that was not used for training and testing (unseen individuals, N = 152,733). This cohort contained 18,537 healthy individuals and 134,196 individuals with neurological and psychiatric conditions (as defined by the ICD-9 and ICD-10 codes described before). Age bias correction was performed in the testing and independent validation dataset by utilizing the beta and alpha parameters fitted in the training cohort.

#### Comparison with PhenoAge

We retrieved the following blood biomarkers from EHR in the All of Us Cohort: Albumin, creatinine, glucose, C-reactive protein, lymphocyte count, MCV, RDW, white cells blood count, and alkaline phosphatase. We only kept individuals with all biomarkers available (N=8,386). We then estimated PhenoAge, using the coefficients and equations provided in (11). We evaluated the performance of PhenoAge using metrics described previously. In addition, we computed the correlation between PhenoAge and ClinAGE.

#### Genetic ancestry estimation

genetic ancestry groups were predicted by the Data and Research Center (DRC) with a random forest classifier trained on 16 genetic principal components (PCs), using the 1000 genomes project and Human Genome Diversity Project (HGDP) as reference (61). In detail, shared sites between the All of Us cohort and the reference panel (1KG and HGDP) were used. Only autosomal and biallelic SNPs with a frequency > 0.1% and a call rate > 99% were kept. An R^2^ = 0.1 was used to determine variants in linkage disequilibrium, keeping 130,660 independent SNPs. Genetic PCs were then derived using the hwe_normalized_pca function in the Hail Python package. After, a random forest classifier was trained on the reference panel using this set of high-quality variants. All individuals were assigned to an ancestral group with a certain probability. However, to diminish population structure, we selected individuals with a posterior probability of belonging to their assigned group > 0.75, keeping a total of 120,761 individuals for analysis.

### Association with age-related traits

#### Laboratory measurements

We retrieved 89 laboratory measurements collected from EHR, available through All of Us CDR v8. We selected markers that were measured within one year of recruitment, as explained for the biomarkers used to train the model. These age-related biomarkers included: Protein (urine), Glucose (urine), Hemoglobin (urine), Ketones, Indirect bilirubin, Albumin/Globulin ratio, Hemoglobin A1c, Globulin and estimated Glomerular filtration rate, Iron (blood), among others (89 in total, Supplementary Table 3). We then scaled every measurement to mean 0 and variance 1 using the scaling function in the scikit-learn package in Python. Association between ClinBAG and these biomarkers was tested by linear regression, adjusted by sex, chronological age and the first 10 genetic PCs. To decrease the impact of reverse causation on the analysis, we performed a second association but using the same makers measured within one year of the last follow-up (n=92). We evaluated if the significant associations were maintained at this second time-point, and we selected those as the ones strongly associated with our biological age model. All p-values were adjusted for multiple testing using FDR correction with the Benjamini-Hochberg method.

#### Sociodemographic and self-reported variables

We retrieved questionnaire data from All of Us, including education level, annual income, health insurance, physical health perception, difficulty walking or climbing and difficulty concentrating, as defined by the enrollment survey that All of Us performs for all participants. We binarized variables with multiple categories in the following way: Low income was defined as an annual income <75,000 USD, low education was defined as less than grade 12^th^, as suggested by Nakao et al. (26). For self-reported physical health, we aggregated “good”, “very good” and “excellent” into a good health category and “poor” and “fair” as “poor health”. For difficulty concentrating and climbing/walking, we selected individuals who answered “yes” and “no” and removed missing values.

#### Cognitive variables

We retrieved data from two cognitive tests available in All of Us: Guess the Emotion, a facial emotion recognition task where participants are presented with images of people and asked to identify their emotion based on facial expression. We analyzed mean reaction times to determine the correct emotion (sad, fearful, angry and happy). We analyzed these reaction times by emotion and in aggregate, as well as overall accuracy (0-100) and a score assigned by the test based on correct answers. We excluded tests where the median response time for correct answers (medianRTc) is less than 700 ms (flag_medianRT=1), and where participants responded to the same option more than 90% of the trials (flag_sameResponse=1), and participants for whom >10% of trials had a reaction time less than 300 ms (flag_trialFlags=1).

City or Mountain test: classified as a gradual onset continuous performance task (gradCPT), it aims to test how well participants resist responding to a changing scene. Participants are asked to press a button when they see an image of a city while fading images of cities or mountains appear on the screen. This test is used to understand attention, cognitive control and the ability to respond to information while ignoring distractions. Quality control included removing participants with the following flags: Flag_trailFlags = 1: if all test trials were longer than 60 seconds, suggesting that the participant was taking breaks. Flag_nonResponse =1: if at any point the participant withheld their response for 75 consecutive test trials (60 seconds). Flag_omissionErrorRate=1: when a participant withholds their response on more than 50% of go test trials (city images).

### Phenotype associations

We retrieved phenotype information for the independent cohort (N = 152,733) by matching ICD-9 codes from 290-359 and ICD-10-F and G codes curated from AoU EHR, as described before (26). We matched these ICD codes to phecodes, defined by All of Us, to aggregate different codes into phenotypes. If an individual has at least one ICD-9/ICD-10 code recorded in their EHR, the subject was classified as a case. If none of these codes were recorded, the individual would be classified as a control. This resulted in the definition of 434 phenotypes. If the number of cases was greater than 100, association with ClinBAG was tested by logistic regression, adjusted by sex, chronological age and the first 10 genetic PCs. To account for multiple testing, p-values were corrected using the false discovery rate (FDR), and significance was defined as a p-corrected value < 0.05.

In addition, we selected 190 phenotypes that were significantly associated with ClinBAG for ancestry-stratified analysis. We stratified based on genetic ancestry for the three major groups present in this cohort: European (EUR), admixed Americans (AMR) and Africans (AFR). We performed an association between ClinBAG and a phenotype within each ancestral group if that ancestral group contained at least 50 cases. We adjusted for the same covariates as before, including the first 10 PCs, to account for within-ancestry variation. In addition, we utilized a Cochran Q test to determine if there was heterogeneity between ancestries for each phenotype. All the p-values were corrected for multiple testing using the FDR approach.

Finally, we performed a Cox regression analysis using incident cases. We prioritized the most significant phenotypes at the cross-sectional level and the ones that showed heterogeneity between ancestries. We defined the start of the follow-up time as one year after the recruitment date, to ensure that the biomarkers used to train the biological age model were measured before the start of the follow-up. Individuals at risk were defined as individuals with no neurological phenotypes at baseline (using ICD codes) and incident cases for every phenotype. In addition, we utilized the last visit date as the end of the follow-up. This Cox regression was adjusted by the same covariates as the cross-sectional model. Moreover, we visualized this time to event outcomes using a Kaplan-Meier plot, by dividing every cohort by the median into high and low ClinBAG.

### Sample and variant quality control for WGS

We retrieved pfiles (PLINK2) generated from WGS, located in the All of Us workbench CDR v.8 (ACAF pfiles), that include variants with an allele frequency of greater than 1% in at least one specific population. We restricted these files to individuals from our independent cohort using the –keep flag in PLINK2 (n = 120,761) (63). We excluded individuals flagged as bad quality by All of Us quality control of VCFs, meaning that they failed any QC metric from GATK after variant calling. We then excluded individuals with genetically undetermined sex and discordant sex. After, we removed variants with less than 20 allele counts (--mac 20), variants failing the Hardy-Weinberg equilibrium (1x10^−6^), and with more than 10% of missing genotypes (--geno 0.1) within each ancestry group (AFR, EUR, AMR and EAS). Finally, variants were pruned based on LD (PLINK2 parameter --indep-pairwise 1000 50 0.2), resulting in around 400k-800k variants, depending on the ancestry (higher number for AFR).

### Common variants genome-wide association study (GWAS)

We conducted a GWAS including related individuals with REGENIE 2.02 (64). Briefly, the pruned pfile was used as an input for REGENIE step 1, using ClinBAG as phenotype, sex, the first 10 PCs and sequencing site as covariates for each ancestry group separately. We used a block size of 1000 for LD computation. For REGENIE step 2, we utilized all variants without pruning on the same phenotype and covariates.

### LD score regression

Genetic correlations between BAG GWAS between ancestries were computed using LDSC (65). In addition, we estimated the LD regression intercept, lambda GC and ratio to evaluate inflation and confounding. LD scores utilized as reference were retrieved from the UK biobank for each ancestry, as described elsewhere (66).

### Heritability estimation

We estimated the biological age gap heritability using GCTA GREML (GCTA version 1.95.0) (67) in 10,000 unrelated samples for each ancestry group with variants with an allele frequency > 1%. The model was adjusted by the same covariates as the GWAS.

#### Fine mapping

To identify the most likely causal variant for each genomic risk locus for ClinBAG GWAS in each ancestry, we performed fine mapping with Susie (68). We first identified the lead variant for each locus and expanded 1 Mb on each side of this variant. We then estimated the ancestry-specific linkage disequilibrium matrix for variants included in that window using the –r square flag in PLINK1.9 in the cohort data. We identified 95% credible sets using this LD matrix and GWAS summary statistics with Susie.

### Variant annotation and enrichment

Summary statistics were annotated using FUMA (69). Genome-wide significance was defined as P < 5x10^-8^, and independent risk loci were defined based on FUMA’s SNP2GENE module (find version). SNP2GENE groups variants into genomic risk loci using LD structures from the 1000 Genomes Project. For each ancestry, we selected the appropriate LD reference panel (AFR, EUR, AMR) and a threshold of R^2^ of 0.6 (default FUMA parameters). For annotation of genomic loci, we used the lead SNP, reported: 1) the closest gene (based on distance between lead SNP and transcription start site), 2) genes in LD (based on positional location in regions with variants with R2 > 0.2 with the lead variants and 3) eGenes (based on GTEx v10 with a genome wide significant tissue eQTL < 5x10^-8^). In addition, we functionally annotated variants using ANNOVAR, as implemented in FUMA. We prioritized variants in coding regions and with moderate impact at the protein level (CADD score > 7). For variants located in introns or intergenic regions, we prioritized using eQTLs. We added annotations from the GWAS catalogue to identify variants previously associated with other traits, and we checked the ancestry of such GWAS results to assess consistency.

### Gene-level annotation and enrichment analysis

We used MAGMA v1.08 (70) as implemented in FUMA (69) to test associations between SNPs and genes to generate gene-level association statistics (SNP2GENE module). All analyses were performed in FUMA using default parameters: First, SNPs were assigned to genes using Ensembl v102 gene boundaries (excluding the extended MHC region between 25Mb-33Mb). Gene-level testing was performed based on GWAS summary statistics of ClinBAG by ancestry, and association P-values were corrected for the number of tested genes (n=18823 protein coding genes, significant threshold 2x10^-6^). Resulting associated genes were used as input for enrichment analysis in several databases, including Gene Ontology Biological Processes (GOBP), Reactome, GWAScatalog, KEGG, Wikipathways, Oncogenic signatures, Hallmark gene sets and BioCarta. We included 54 different tissues from GTEx v8. Significance was defined as a false discovery rate (FDR) < 0.05.

### scDRS analysis

Single-cell Disease relevance score (scDRS) analysis was performed with scDRS v1.03 (71). This method identifies cell types within a single-cell RNA sequencing dataset that overexpressed genes from the ClinBAG GWAS that were prioritized by MAGMA (70). We downloaded a single-cell RNA-seq dataset from peripheral blood mononuclear cells (PBMCs, N = 1.9M) (17) from different age groups: A (25-34), B (35-44), C (45-54), D (55-64), E(65+), obtained from healthy individuals. Briefly, this cohort includes European, non-obese (BMI < 30) males and females. Inclusion criteria included: non-smokers, no history of cancer or inflammatory conditions such as arthritis, Crohn’s disease, colitis, dermatitis, or lupus or blood-borne infections such as HIV, hepatitis B-C (17). Samples were sequenced using 10X Chromium technology. We obtained log-normalized counts, barcodes and associated metadata (including cell type annotation) from the Synapse database (syn49637038). Samples were integrated using Harmony (72) (as implemented in harmonypy) to account for batch effects. The UMAP was computed using the first 30 principal components. We stratified the h5ad file by age group using Scanpy XX (73). The top 1000 ClinBAG GWAS genes and their Z-scores as weights for each ancestry (EUR and AFR) generated using the MAGMA gene-based association analysis were used as input for scDRS. The scDRS compute_score command was run with default parameters for each ancestry with the top 1000 genes from MAGMA as gene set, and 9 cell types: B cells, CD4+ T cells, Double negative T cells (DN T cells), MAIT cells, Myeloid cells, Natural Killer cells (NK cells), Progenitor cells, TRAV1-2-CD8+ T cells and gamma-delta T cells (gd T cells). After stratifying by age group, every cell type contained >100 cells. scDRS perform_downstream command was used with default parameters to obtain the enrichment of each gene set per cell type.

### Sickle cell anemia variant rs334 analysis

To further explore the association between rs334 and the biological age gap in Africans, we identified homozygous individuals for the reference allele TT (N=20314), heterozygous AT (N=1926), and homozygous AA (N=71), who have sickle cell anemia disease. We then compared the biological age gap, using individuals with genotype TT as reference, and adjusting for sex, age, and 10 genetic PCs. We repeated the analysis, but comparing each of the 22 features used to train the model to identify a specific feature that could explain the patterns observed in the biological age gap distribution.

### NPRL3 expression analysis

To investigate specific cell types that express NPRL3 in the context of our biological aging clock, we retrieved single-cell RNA-seq data from bone marrow and PBMCs from healthy individuals in the GEO database (GSE253355) (23). Cell annotation was retrieved from the Seurat file, containing 35 cell types as described elsewhere (23). We visualized the log normalized expression of NPRL3 using the dotplot and violin plot functions, as implemented in Seurat V5.

## Data availability

The genotypes and phenotypes of All of Us participants are available through application to the All of Us research program (https://allofus.nih.gov/). ClinBAG GWAS summary statistics will be available upon publication

## Code availability

The code used to perform the analyses is available on GitHub upon publication

## Author contributions

Conception and design: A.M-G, A.L and S.Z. Formal analysis: A.M-G, C.Y. S., T.M-Z, H.M.T, A.R. Result interpretation and visualization: A.M-G, A.L, S.Z, C.Y. S., T.M-Z, H.M.T, D.H, S.Y, G.L, V.M. Writing – review and editing: A.M-G, A.L, S.Z, C.Y. S., T.M-Z, H.M.T, D.H, S.Y, G.L, V.M. Supervision: A.L and S.Z. All authors revised and approved the final version of the manuscript.

## Competing interests

Authors have declared no competing interests.

## Acknowledgments

We would like to thank the participants who provided biological samples and data for All of Us. A.M-G is supported by the Stage Quebec program and the Canadian Institute for health Research (CIHR) doctoral scholarship, Funding reference number: 210230. S.Z. is supported by a Fonds de recherche du Québec santé (FRQS) Chercheurs-boursiers J1 award. This research was supported by the Canadian Institute for Health Research (CIHR) project grant PJT205932 (S.Z).

## Notes

### Competing Interest Statement

The authors have declared no competing interest.

### Author Declarations

Ethics committee/IRBof McGill University gave ethical approval for this work, granting access to All of Us biobank (IRB: 2107073).

